# Quantifying the vaccine-induced humoral immune response to spike-receptor binding domain as a surrogate for neutralization testing following mRNA-1273 (Spikevax) vaccination against COVID-19

**DOI:** 10.1101/2022.03.09.22271896

**Authors:** Imke Kirste, Sayuri Hortsch, Veit Peter Grunert, Holly Legault, Maha Maglinao, Udo Eichenlaub, Basel Kashlan, Rolando Pajon, Simon Jochum

## Abstract

**Background:** There is a need for automated, high throughput assays to quantify immune response after vaccination against severe acute respiratory syndrome coronavirus 2 (SARS-CoV-2). This study assessed the combined utility of the Roche assays, Elecsys^®^ Anti-SARS-CoV-2 S (ACOV2S) and Elecsys Anti-SARS-CoV-2 (ACOV2N) using samples from the 2019-nCoV vaccine (mRNA-1273, Spikevax™) phase 2 trial (NCT04405076).

**Methods:** Samples from 593 healthy participants in two age cohorts (18–54 years and ≥55 years), who received two injections with either placebo (n=198) or mRNA-1273 at a dose of either 50 μg (n=197) or 100 μg (n=198), were collected at Days 1 (first vaccination), 15, 29 (second vaccination), 43 and 57. ACOV2S results were used to assess the humoral response to vaccination in different clinical trial subgroups and were compared to a live virus microneutralization assay. Sample panels from patients with evidence of previous or concomitant infection (as identified using ACOV2N) or with an inconsistent antibody response pattern were analyzed separately.

**Results:** Receptor-binding domain (RBD)-specific antibodies were readily detectable by ACOV2S for the vast majority of participants (174/189 [50 μg dose group] and 178/192 [100 μg]) at the first time point of assessment, with non-converters predominantly older in age. Complete seroconversion for all participants was observed at the subsequent timepoint (Day 29) and before administration of the second dose of vaccine. Two weeks after the first vaccine dose (Day 15), geometric mean concentration (GMC) of antibody levels were 1.37-fold higher in the 100 μg compared with the 50 μg dose group; this difference reduced to 1.09-fold two weeks after the second dose (Day 43). In both the 50 μg and 100 μg dose groups, a more pronounced response was observed in the younger versus the older age group on Day 15 (2.49-fold and 3.94-fold higher GMC, respectively) and Day 43 (1.35-fold and 1.50-fold higher GMC). Few subjects had a previous or concomitant natural SARS-CoV-2-infection (n=8). Vaccination of pre-infected individuals boosted the immune response to very high ACOV2S results compared to infection-naïve vaccine recipients. ACOV2S measurements were strongly correlated with those from the live microneutralization assay (Pearson’s r=0.779; p<0.0001) and good qualitative agreement was achieved (100% positive and 91.8% negative percentage agreement; 90.0% positive and 100% negative predictive value).

**Conclusion:** The results from this study confirmed that ACOV2S is a highly valuable assay for the tracking of vaccine-related immune responses. Combined application with ACOV2N enables serologic monitoring for breakthrough infection or stratification of previous natively-infected individuals. The adaptive measuring range and high resolution of ACOV2S allows for the early identification of seroconversion as well as for resolution of very high titers and detection of longitudinal differences between age and dose groups. Additionally, good correlation of ACOV2S with live virus microneutralization indicates the utility of ACOV2S as a reliable estimate of neutralization capacity in routine diagnostic settings.

## 1. Introduction

Severe acute respiratory syndrome coronavirus 2 (SARS-CoV-2) is a highly transmissible and pathogenic coronavirus that has infected hundreds of millions of people globally since it first emerged in 2019 [1]. To reduce the burden of disease, vaccines have been rapidly developed and administered extensively worldwide [2]. While vaccines can be directed against all viral SARS-CoV-2 proteins, [3, 4] the spike (S) and nucleocapsid (N) proteins are considered the main targets of immune response. To date, the majority of approved vaccines and vaccine candidates are targeted to the S antigen, which facilitates entry to host cells via the angiotensin-converting enzyme 2 (ACE 2) receptor [5]. The immunogenicity and functional importance for host cell entry of the S antigen render it an attractive target for vaccination with strong neutralizing potential. However, strong antigenic drift and immune evasion by the virus was expected and has already occurred in emerging strains of SARS-CoV-2 [6].

There is growing evidence that the presence of neutralizing antibodies and immunoglobulins can be correlated with protection against SARS-CoV-2 [3, 4, 7, 8]. Although the measurement of neutralizing antibodies by live virus neutralization assays provides a means of direct functional assessment, the requirement for cell culture and live virus preparations renders robust standardization challenging even when applying the same technical setup. As a surrogate for neutralizing titers, the quantification of antibody concentrations using commercially-available immunoassays is an attractive option for measuring the response to vaccination and is supported by observations of a correlation between antibodies targeting the receptor-binding domain (RBD) and virus neutralizing titers in plasma from patients naturally infected with SARS-CoV-2 [9-14].

Recent publications have shown that immune response to vaccination is dependent on vaccine type, age, and comorbidities [15-19] and debates have arisen regarding the need to provide additional doses to vulnerable populations [19, 20]. More recently, the emergence of highly transmissible variants, such as Omicron, has led to a number of countries offering booster vaccination doses to the general population [21], with boosters now known to produce a significant increase in neutralizing activity against the Omicron variant [22]. To further understand the potential benefits of offering additional vaccine doses, there is a high need for sensitive and specific assays that can reliably quantify immune responses to vaccination. In addition, large data sets from well-controlled studies are needed to generate the best possible estimates on antibody response to vaccination.

The Roche Elecsys^®^ Anti-SARS-CoV-2 S assay (hereby referred to as ACOV2S) is an automated, high throughput assay that quantifies antibodies against the RBD of the S protein, developed to detect low levels of such antibodies with high sensitivity (97.92%) and specificity (99.95%) [23]. In contrast, the Roche Elecsys Anti-SARS-CoV-2 immunoassay (hereby referred to as ACOV2N) specifically identifies antibodies to the N protein and therefore can only detect humoral responses elicited following natural infection and not vaccines targeting the S protein [24]. In a recent analysis of samples from a phase 1 trial of the mRNA-1273 vaccine (Spikevax^TM;^ Moderna, Cambridge, MA), ACOV2S showed absence of any baseline noise indicating high specificity for detecting a vaccine-induced immune response [25]. In addition, moderate to strong correlations were observed between ACOV2S and several neutralization assays.

To generate further evidence of the clinical utility of ACOV2S and ACOV2N, we utilized samples from participants enrolled in a phase 2 trial of the mRNA-1273 vaccine. This study provided a large sample size to confirm previous observations obtained using the phase 1 dataset. In addition, the inclusion of a placebo arm in this study provided additional evidence of the specificity of ACOV2S for the detection of a vaccine-induced immune response. Furthermore, the study included participants in two age groups (18–54 years and ≥ 55 years), allowing us to determine whether ACOV2S is capable of detecting age-related differences in humoral response. We also compared the results from ACOV2S with results from a live virus neutralization assay used in the phase 2 study. Finally, the combined use of ACOV2S and ACOV2N provided a method to discriminate between immune responses after vaccination in individuals with or without previous or concomitant SARS-CoV-2 infection.

## 2. Methods

### 2.1. Study design and participants

In this retrospective exploratory analysis, stored samples from participants enrolled in the phase 2 trial of mRNA-1273 (NCT04405076) were included for assessment. Full methodological details of this study, including collection of blood samples, have been described previously [26]. In brief, healthy participants aged ≥18 years in two age cohorts (aged 18-54 years and aged ≥55 years) were randomized 1:1:1 to receive 50 μg or 100 μg of mRNA-1273 or placebo. The vaccine and placebo were administered using a 2-dose regimen with the first dose given on Day 1 and the second on Day 29. All participants were screened and randomized between May 22 and July 8, 2020. Informed written consent was originally obtained from all study participants in the context of the associated vaccine phase 2 study and the study was conducted in accordance with the principles of the Declaration of Helsinki and Good Clinical Practice Guidelines. Approval was granted by the regulatory and institutional committees for the phase 2 trial [26] and the diagnostic protocol under which the existing samples were tested.

Blood samples were collected at baseline (Day 1, first vaccination), and Days 15, 29 (second vaccination), 43, and 57, were analyzed and serum testing was performed at PPD central laboratory (Highland Heights, KY, USA).

### 2.2. Laboratory assays

#### 2.2.1. Elecsys Anti-SARS-CoV-2-S (ACOV2S) immunoassay

The ACOV2S assay has been described previously [25]. In brief, samples were quantified for SARS-CoV-2 RBD antibodies with a measuring range of 0.4-25,000 U/mL The assigned U/mL are equivalent to Binding Antibody Units (BAU)/mL as defined by the first World Health Organization (WHO) International Standard for anti-SARS-CoV-2 immunoglobulin (NIBSC code 20/136). Values that deviated by more than three times the interquartile range (IQR) from the lower or upper quartile of the assay results were defined as statistical outliers. All outliers were associated with inconsistent patterns of antibody titers, most likely originating from sample mis assignments rather than biological or technical effects.

#### 2.2.2. Elecsys Anti-SARS-CoV-2 (ACOV2N) immunoassay

In addition to the quantification of RBD-specific antibody titers induced by mRNA-1273 vaccination, all samples were assessed on the same cobas e 602 module with the ACOV2N assay [24]. As natural infection with SARS-CoV-2 but not vaccination with mRNA-1273 can trigger a positive ACOV2N result, this assay was used to determine whether participants had been infected naturally with SARS-CoV-2 either before or during the period of investigation.

#### 2.2.3. Comparator assays

Neutralizing antibody levels were determined under the phase 2 study protocol [26] and the results were transferred to Roche for analysis. Serum neutralizing antibody titers against SARS-CoV-2 were measured using a live virus microneutralization (MN) assay based on an *in situ* ELISA readout; further details of this assay can be found in the supplementary appendix of the phase 2 publication [26]. The final reportable value for each sample was the MN_50_ titer which refers to the dilution required to achieve 50% neutralization.

In case no significant inhibition of infection was observed (<50% neutralization) with the MN assay, the assay result was qualitatively interpreted as negative for neutralizing activity in all qualitative concordance analyses. In this case, the missing quantitative result was substituted by half the lower limit of quantitation. Samples showing significant inhibition (≥50% neutralization) at any of the applied dilutions were interpreted as positive for neutralizing activity in all qualitative concordance analyses. In case the titer exceeded the measuring range of the MN assay, the quantitative result was substituted by the upper limit of quantitation. Statistical outliers were defined as values that deviated by more than three times the interquartile range (IQR) from the lower or upper quartile of the assay results were defined as statistical outliers. Of note, most MN_50_ titers from participants randomized into one of the vaccine groups exceeded the assay’s measuring range at later visits (Days 43 and 57), so the IQR was severely underestimated and the criterion could not be applied to those visits. A Gaussian distribution fit to the right-censored data with subsequent application of the 3xIQR criterion did not detect any further outliers.

### 2.3. Statistical analysis

To evaluate the humoral response to vaccination with mRNA-1273, analyses were performed on samples from SARS-CoV-2-naïve participants. Statistical ACOV2S outliers were described separately and were also included in a sensitivity analysis. Confidence in analyzing outliers separately was based on the previously observed reliable performance of ACOV2S [23] and on immunobiological rationale [25]. For the comparison of ACOV2S and the live MN_50_ assay, the analysis was performed using samples from SARS-CoV-2-naïve participants, excluding those with either an ACOV2S or MN_50_ outlier result.

For each age category and dosage group, ACOV2S-measured anti-RBD antibody levels are shown as line plots and boxplots (log-scale) for every measurement time point. Comparison of ACOV2S-measured antibody levels per age category, dose group, and time point were conducted using reverse cumulative distribution curves. For ACOV2S, geometric mean concentrations (GMCs) and geometric mean fold rises (GMFRs) were calculated for each time point and stratified by age category and dose group. 95% confidence intervals (CIs) were calculated by Student’s t distribution on log-transformed data and subsequent back-transformation to original scale. For the assessment of seroconversion, as measured by ACOV2S, the percentage of subjects who crossed the reactivity cut-off at 0.8 U/mL at or before a given time point was evaluated. For all analyses, values below the measuring range were set to the numeric value of 0.4 U/mL and values above the measuring range were set to 25,000 U/mL.

To assess the concordance of ACOV2S with the live virus MN_50_ assay, a pairwise method comparison across all available data points (excluding values outside the measuring range) using Passing-Bablok (log-scale) regression analyses [27] with 95% bootstrap CIs were provided and Pearson’s correlation coefficients (r) with 95% CIs were calculated. A comparison of the geometric mean fold rises (GMFRs) was also performed to compare the dynamic range of the ACOV2S and MN assays.

Qualitative agreement between ACOV2S and the MN_50_ assay was analyzed by positive percentage agreement (PPA), negative percentage agreement (NPA), and overall percentage agreement (OPA), positive and negative predictive value (PPV and NPV) with exact 95% binomial CIs, and the positive and negative likelihood ratio with 95% CIs calculated (per Simel et al. approximation [27]). The software R, version 3.4.0, was used for statistical analysis and visualization [28].

## 3. Results

This analysis included longitudinal sample panels from a total of 593 participants; of these, 295 were aged 18-54 years and 298 were aged ≥55 years. Of the overall population, 198 participants received placebo, 197 received the 50 μg dose of mRNA-1273 and 198 received the 100 μg dose of mRNA-1273. In the placebo, 50 μg, and 100 μg dose groups, respectively, mean ages (±SD) were 51.1 (±15.6), 50.6 (±16.2), and 51.4 (±15.3) years and 35.4%, 30.5%, and 37.9% were males. In total, 15 subjects out of 593 were excluded from analysis including eight participants with at least one positive ACOV2N result (five were positive at baseline and three tested positive for ACOV2N during the course of the study) and seven participants identified as statistical outliers (six for ACOV2S and one for the live MN_50_ assay).

### 3.1. Humoral response after vaccination with mRNA-1273 assessed by Elecsys Anti-SARS-CoV-2 S assay

After vaccination, anti-RBD antibody levels tended to increase until Day 43 for both dose groups before dropping slightly by Day 57 (Figures 1A and 2A and Supplementary Table 1). RBD-specific antibody levels indicated that almost all vaccinated participants had seroconverted prior to Day 15 (Supplementary Table 1; 50 μg: 174/189 [92.1%], 100 μg: 178/192 [92.7%]). Rates of seroconversion at Day 15 were slightly higher for participants aged 18-54 years compared with participants aged ≥55 years in both dose groups (50 μg: 96.8% vs 87.5%; 100 μg: 97.9% vs 87.6%). In addition, patients who had not seroconverted by Day 15 tended to be older than the overall patient populations in the two vaccine arms (median [IQR] age: 66.0 years [58.0–72.0] vs 55.0 years [38.0–64.0]). By Day 29, i.e., day of second vaccination, all vaccinated participants had seroconverted. For both dose groups, GMCs were notably higher two weeks after the second vaccine dose (Day 43; 50 μg: 6980 U/L, 100 μg: 7638 U/L) compared with Day 29 (50 μg: 88.7 U/L, 100 μg: 117 U/L) (Supplementary Table 1). Overall, 12 out of 2838 samples had measured antibody levels that exceeded the applied upper limit of quantitation of 25,000 U/mL, all of which were taken after the second vaccination. The distribution of the ACOV2S levels after the first vaccine dose appear more heterogeneous than after the second vaccine dose (Figure 1B). In addition, there is greater heterogeneity in the older age group compared to the younger age group.

**Figure 1.**
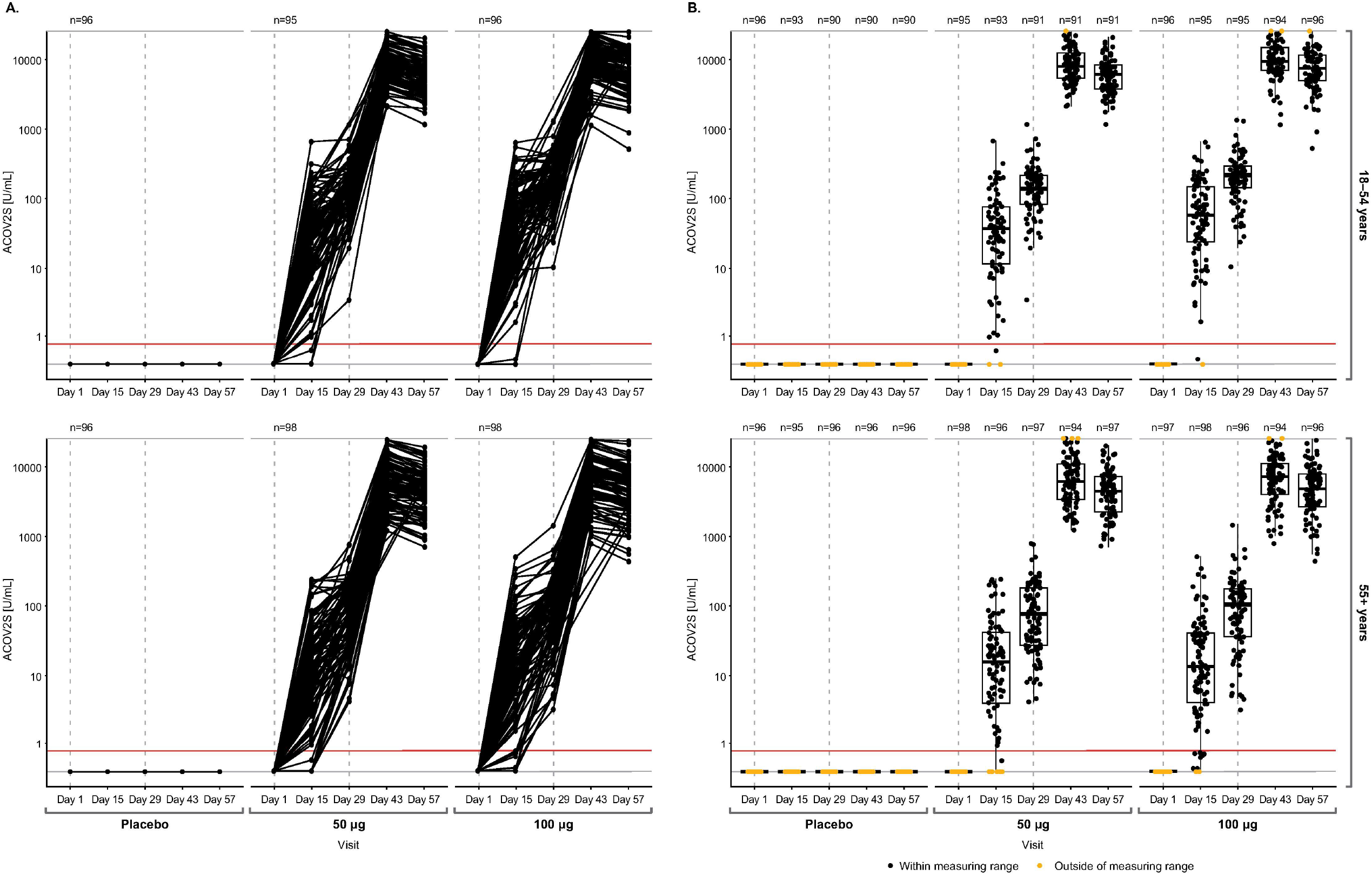
Time course of ACOV2S-measured antibody levels following mRNA-1273 (Spikevax) vaccination. Antibody levels following vaccination are shown as line plots in Panel A and box plots in Panel B (top and bottom panels showing results from 18–54 and 55+ years age groups, respectively). Dotted grey vertical lines indicate time of vaccination, administered at Days 1 and 29. Box plots show the individual readouts (black dots) and, 25^th^, 50^th^, and 75^th^ percentiles (black box). Red horizontal line indicates reactivity cut-off (0.8 U/mL). Values below the measuring range were set to the numeric value of 0.4 U/mL and values above the measuring set to 25,000 U/mL (yellow dots).

**Figure 2.**
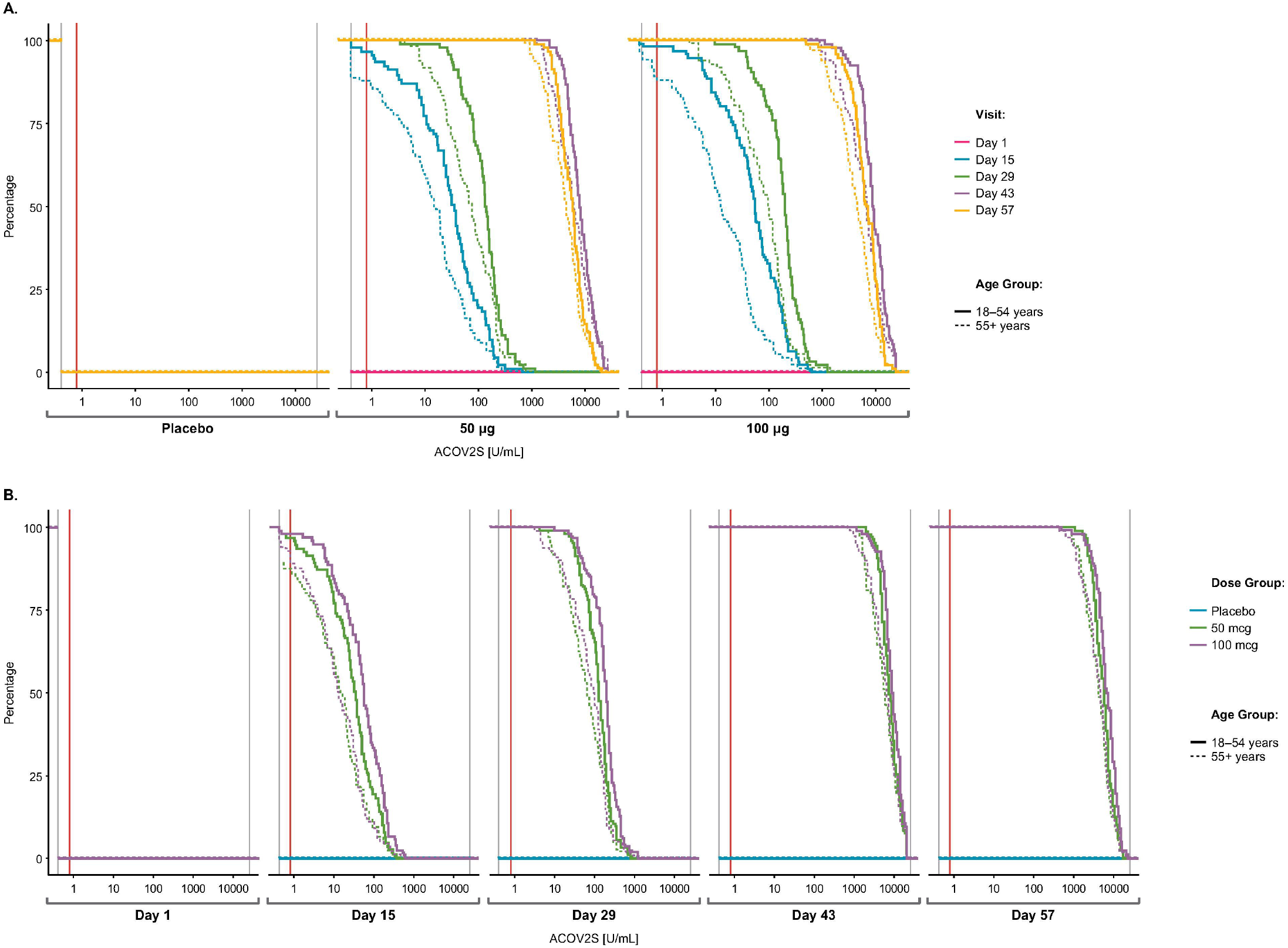
Time-dependent antibody responses as measured by ACOV2S following mRNA-1273 (Spikevax) vaccination. Reverse cumulative distribution curves allow for comparison of ACOV2S-measured antibody level distributions between subgroups (Panel A) and visits (panel B). Red vertical line indicates reactivity cut off (0.8 U/mL).

The determined antibody concentrations correlated with the applied vaccine in a dose-dependent manner (Figure 2), with a GMR point estimate between the 100 μg versus the 50 μg group above 1 at follow-up visits (Supplementary Table 1). Differences between the 100 μg and 50 μg dose groups were more prominent after the first vaccination (Day 15, 1.37-fold; Day 29, 1.32-fold higher) than after the second vaccination (Day 43, 1.09-fold; Day 57, 1.12-fold higher). A more pronounced response was also observed in the younger compared to the older age groups (Figure 2B) in both the 50 μg dose group (Day 15, 2.49-fold; Day 29, 1.93-fold; Day 43, 1.35-fold; Day 57, 1.35-fold higher) and in the 100 μg dose group (Day 15, 3.94-fold; Day 29, 2.44-fold; Day 43, 1.50-fold; Day 57, 1.57-fold higher).

Of the eight participants with at least one positive ACOV2N result during the trial indicative of a natural SARS-CoV-2 infection, two were in the placebo group and both showed a modest increase in both ACOV2N and ACOV2S levels between Days 43 and 57. A further three participants demonstrated a highly robust response to the mRNA-1273 vaccine, either reaching or exceeding the upper limit of quantitation for the ACOV2S assay. Line plots for the ACOV2S results for these eight participants are shown in Supplementary Figure 1.

Line plots of the ACOV2S values of a further six participants with ACOV2S outliers are shown in Supplementary Figure 2. Performed for completeness, a sensitivity analysis including these outliers did not significantly change the outcome of the analysis of humoral response to vaccination with the mRNA-1273 vaccine (Supplementary Figures 3 and 4).

### 3.2. Concordance of ACOV2S with the live microneutralization assay

Figure 3 visualizes concordance of ACOV2S with the live MN_50_ assay. Good numerical correlation was observed (Pearson’s r=0.779 [95% CI 0.742–0.811]; p<0.0001; determined on log-log scale). Of the 992 samples that were reactive in the ACOV2S assay, the majority (893 samples) exhibited significant *in vitro* neutralization capacity. The remaining 99 samples that did not yet show *in vitro* neutralization capacity were taken before the second vaccination and were of relatively low titer. Of these, 43.4% (43/99) were from the younger age group and 41.4 % (41/99) were from the 100 μg dose group. The ACOV2S GMC in these 99 samples was 37.0 U/mL (65.9 U/mL in the 18-54 age group, 23.7 U/mL in the ≥55 age group), and thus significantly lower than the overall GMC of 102 U/mL in the two groups at Day 29. Qualitative agreement between ACOV2S and MN_50_ results is presented in Table 1. Using the 0.8 U/mL ACOV2S cutoff, the PPA and NPV were 100%. The NPA was 91.8%, due to the samples which were already reactive for ACOV2S but did not yet show neutralization in the MN_50_ method. The NPA was modestly increased to 93.4% if a higher ACOV2S cutoff of 15 U/mL was used; this higher cutoff has been observed to correlate better with neutralization in samples from participants with a natural infection [29]. A comparison of GMFRs over time (Figure 4) demonstrated higher MN_50_ values in patients aged 18L54 vs ≥55 years and with the 100 vs 50 μg dose, but these differences diminish after the second vaccine dose. These observations are in line with ACOV2S GMFRs, although the ability of the GMFRs using the MN_50_ method to resolve age-dependent effects is less pronounced than using the ACOV2S method. Of note, the measuring range of the MN_50_ assay is limited and most titers exceed the range after Day 29. This limitation contributes to the impaired differentiation by MN_50_ at later timepoints and biases the value of GMFRs.

**Table 1.** Summary of qualitative agreement measures between Elecsys ACOV2S and live microneutralization assays (MN_50_ endpoint; reference) with modified ACOV2S cutoffs.

|  | **0.8 U/mL ACOV2S cut off** | **15 U/mL ACOV2S cut off** |
| --- | --- | --- |
| **PPA** | 100 (99.6–100) | 99.7 (99.0–99.9) |
| **NPA** | 91.8 (90.1–93.3) | 93.4 (91.8–94.7) |
| **OPA** | 95.3 (94.3–96.2) | 96.0 (95.1–96.8) |
| **PPV** | 90.0 (88.0–91.8) | 91.8 (89.8–93.4) |
| **NPV** | 100 (99.7–100) | 99.7 (99.2–99.9) |
| **Positive likelihood ratio (95% CI)** | 12.2 (10.1–14.7) | 15.0 (12.2–18.6) |
| **Negative likelihood ratio (95% CI)** | 0 (0–NA) | 0.00360 (0.00116–0.0111) |

Data shown as % (95% CI) unless otherwise stated.
NPA, negative percent agreement; NPV, negative predictive value; OPA, overall percent agreement; PPA, positive percent agreement; PPV, positive predictive value.

**Figure 3.**
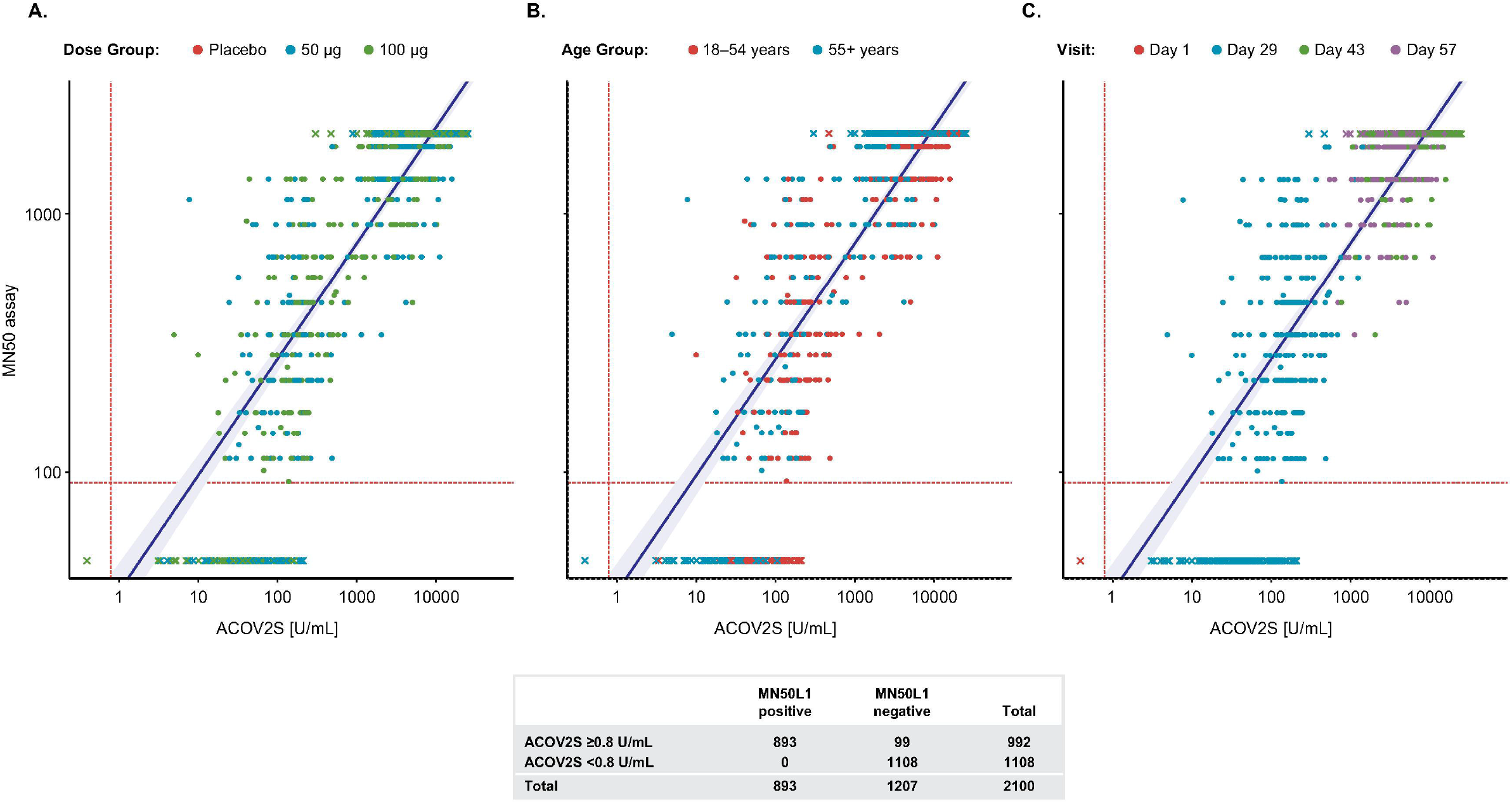
Comparison of ACOV2S and the live microneutralization assay (endpoint MN_50_). Passing-Bablok regression fit (log scale) of ACOV2S with MN assay results (Pearson’s r [95% CI]: 0.779 [0.742–0.811]; p<0.0001). The shaded area represents the 95% confidence interval for the fitted curve. Dots or crosses show individual sample readouts. Crosses indicate samples with at least one assay result outside the measuring range. Results are shown for subgroups defined by dose (panel A), age (panel B) and visit (panel C). Overlaid table shows the qualitative agreement between the Elecsys ACOV2S and the MN_50_ assay.

**Figure 4.**
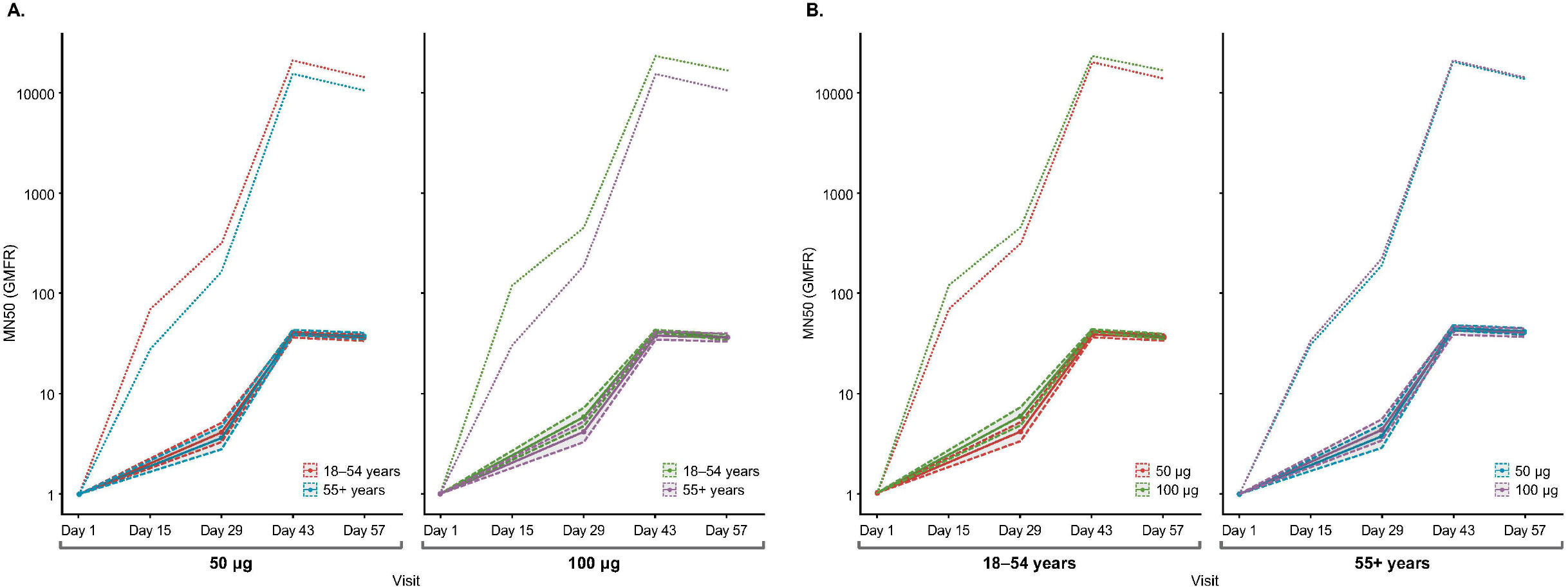
Line plots of MN_50_ GMFRs over time. Comparison of age (panel A) and dose groups (panel B), including 95% CIs. Dotted lines represent the ACOV2S GMFRs.

A sensitivity analysis that included samples from participants with a natural SARS-CoV-2 infection is shown in Supplementary Figure 5 and demonstrated good concordance between the two assays for these samples.

## 4. Discussion

With a rapidly growing number of SARS-CoV-2 vaccines excelling during clinical development, it is essential to reliably quantify the humoral immune response as a prerequisite to determine its value as a marker of response to vaccination and, ideally, as a surrogate marker of the risk of developing symptomatic disease following infection. In this exploratory analysis of mRNA-1273-vaccinated human samples from a phase 2 trial [26], results from the ACOV2S assay demonstrated that anti-RBD antibody concentrations increased in a time-, dose- and age-dependent manner.

Vaccination resulted in seroconversion for 100% of participants within four weeks after the first injection with mRNA-1273, and further large increases in antibody titers were measured after the second injection of the same vaccine irrespective of age or dose group (GMFRs from Day 29 to Day 43 were 78 and 64 in the 50 μg and 100 μg dose groups, respectively). Differences in the immune response after application of a 100 μg vs 50 μg dose were particularly apparent after the first vaccination compared with the second vaccination and also in the younger vs older age groups. A difference in ACOV2S levels between the two age groups was also observed, with higher seroconversion rate and higher ACOV2S concentrations in the younger age group at Day 15. This is consistent with age-dependent antibody responses reported for other COVID-19 vaccines [30] as well as for vaccines for other infectious diseases [31, 32]. The ACOV2S assay has the capability to track immune responses over time with high resolution, which enables differences due to age, dose groups, and other stratification factors to be studied.

In total, eight participants had a positive ACOV2N result either at baseline or at one of the follow-up visits. Of note, no ACOV2N seroconversion was observed after second injection with mRNA-1273 which is in line with reported vaccine efficacy and protection from infection in this phase [26]. Participants who were vaccinated with mRNA-1273 and had a naturally occurring SARS-CoV-2 infection demonstrated titers exceeding those of naïve recipients. This is consistent with a previous study, which reported that individuals with a prior SARS-CoV-2 infection followed by two doses of an mRNA vaccine had higher S antibody measurements compared with individuals with vaccination alone [33]. Presumably, a significantly sustained immune response and most likely increased protection from infection can be expected for these individuals. Comparative long-term monitoring of pre-infected and SARS-CoV-2-naïve individuals will be required to substantiate these assumptions; however, newly emerging SARS-CoV-2 variants with altered transmissibility and virulence add additional levels of complexity. Together, our results further highlight the use of antibody titer assessment after vaccination especially for older subjects with higher variance in response rate. Stronger seroconversion in individuals with prior infection may alleviate the need for a second dose in those individuals, allowing providers to implement a more targeted and individualized approach to vaccination and balancing protection versus possible side effects.

Good correlation was observed between ACOV2S and the live MN_50_ assay. The vast majority of samples with a positive MN_50_ result were also positive by the ACOV2S assay resulting in an excellent PPA and NPV. Some disagreement before second vaccination was noted, where some positive ACOV2S results, albeit of relatively low antibody titer, coincided with non-reactive neutralizing antibody test results. This may be due to higher sensitivity of the ACOV2S assay and the higher quantities of RBD antibodies that may be required for significant *in vitro* neutralization activity [34]. This supports the clinical finding that single-dose vaccination does not convey optimal protection from infection and that the reported antibody titers before the second vaccination are not necessarily indicative of immunity. In contrast, the significantly higher RBD antibody titers following two-step vaccination appear suitable surrogate markers or, to a certain extent, contributing effectors to protection from infection. MN_50_ titers also somewhat reflected age dependency, however, the limitations of the MN_50_ method hampered systematic comparisons, especially at later visits (Days 43 and 57). Concordance was also observed between the ACOV2S and live MN_50_ assays for the samples from participants with a native SARS-CoV-2 infection. This indicates that levels of RBD antibodies are reflective of neutralizing activity irrespective of whether the immune response is induced by infection or vaccination. Consistent with the impact on the ACOV2S results, there is a tendency for participants who had both natural infection and vaccination with mRNA-1273 to have higher MN_50_ values. Overall, the differences in dynamic range and lack of standardization of the MN_50_ assay suggest that the ACOV2S assay is better suited to study the differences across the whole antibody concentration range and all visits.

Overall, these findings are consistent with a previous analysis of ACOV2S values in subjects vaccinated with mRNA-1273 [25] and suggest that ACOV2S-measured antibody levels correlate well with the presence of neutralizing antibodies after vaccination. Combining the high throughput, automated ACOV2S and ACOV2N assays enables stratification of naïve from preinfected individuals and monitoring for concomitant or breakthrough infection. ACOV2S enables reliable quantification of the humoral immune response to vaccination. Furthermore, the observed high titers are in general indicative of convincing in vitro neutralization capacity and hence are very likely associated with immunity that protects from severe disease. Ongoing research is required to elucidate whether anti-RBD thresholds can be defined that are indicative of prevention of symptomatic infection.

Limitations of our study include the relatively short follow-up, which prevents analysis of the ability of the ACOV2S assay to determine the longevity of antibody response. Further comparison studies using longer-term follow-up are warranted. In addition, the analysis is limited to vaccination with mRNA-1273 and may not be generalizable to all SARS-CoV-2 vaccines.

## 5. Conclusion

Using samples from a large phase 2 study, we confirmed previous results from a phase 1 study and demonstrated that the ACOV2S assay can be used to identify and track vaccine-related immune responses. Furthermore, the adaptive dynamic range enabled resolution of a wide titer range and differentiation of the humoral immune response in relation to dose and age. Combination with ACOV2N enabled identification of previous or concomitant natural SARS-CoV-2 infection. Good agreement was observed between ACOV2S and the applied live virus neutralization assay, with discrepancies only observed before the second vaccine dose and in patients with low antibody titers. Additional long-term studies will be required to determine whether the ACOV2S assay can be used to identify patients who may remain at risk of symptomatic infection and may require additional booster vaccinations.

## Data Availability

The authors are committed to sharing data supporting the findings of eligible studies. Access to de-identified patient-level data and supporting clinical documents with qualified external researchers may be available upon request once the trial is complete.

## Acknowledgements

We would like to thank the associated mRNA-1273 Study Team for their contribution to data collection as part of the associated Phase II study (NCT04405076). Additionally, we would like to thank Micah Taylor, Kristin Lucas and the lab operators at PPD Central Labs (Kentucky, US) for their contribution to data collection and study execution. We would also like to acknowledge and thank Celine Leroy, Kristin Nelson, Emma Tao, Yuli Sun, Walter Stoettner, Iliari Layagi (Roche Diagnostics), Laura Schlieker (Staburo GmbH) and Robin Brauckmann (Chrestos Concept GmbH &Co, KG). ELECSYS and COBAS are trademarks of Roche. All other product names and trademarks are the property of their respective owners. The Elecsys Anti-SARS-CoV-2 S assay is approved under Emergency Use Authorisation in the US. Editorial support was provided by Jade Drummond of inScience Communications, Springer Healthcare Ltd, UK, and was funded by Roche Diagnostics International Ltd (Rotkreuz, Switzerland).

## Author contributions

Study concept/design: IK, UE, SJ, SH, RP

Data acquisition: IK, BK

Data analysis and interpretation: SH, VPG, SJ, IK, MM, RP

Review and final approval of manuscript: IK, SH, VPG, HL, MM, BK, UE, RP, SJ

## References

1. Johns Hopkins University Coronavirus Resource Center. COVID-19 dashboard by the Center for Systems Science and Engineering (CSSE) at Johns Hopkins University. 2021 [cited 2021 November]; Available from: https://coronavirus.jhu.edu/map.html.

2. Kyriakidis, N.C., et al., SARS-CoV-2 vaccines strategies: a comprehensive review of phase 3 candidates. NPJ Vaccines, 2021. 6(1): p. 28.

3. Carrillo, J., et al., Humoral immune responses and neutralizing antibodies against SARS-CoV-2; implications in pathogenesis and protective immunity. Biochem Biophys Res Commun, 2021. 538: p. 187–191.

4. Khoury, D.S., et al., Neutralizing antibody levels are highly predictive of immune protection from symptomatic SARS-CoV-2 infection. Nat Med, 2021. 27(7): p. 1205–1211.

5. Dai, L. and G.F. Gao, Viral targets for vaccines against COVID-19. Nat Rev Immunol, 2021. 21(2): p. 73–82.

6. Mengist, H.M., et al., Mutations of SARS-CoV-2 spike protein: Implications on immune evasion and vaccine-induced immunity. Semin Immunol, 2021. 55: p. 101533.

7. Earle, K.A., et al., Evidence for antibody as a protective correlate for COVID-19 vaccines. Vaccine, 2021. 39(32): p. 4423–4428.

8. Gilbert, P.B., et al., Immune correlates analysis of the mRNA-1273 COVID-19 vaccine efficacy clinical trial. Science. 0(0): p. eab3435.

9. Legros, V., et al., A longitudinal study of SARS-CoV-2-infected patients reveals a high correlation between neutralizing antibodies and COVID-19 severity. Cell Mol Immunol, 2021. 18(2): p. 318–327.

10. Salazar, E., et al., Convalescent plasma anti-SARS-CoV-2 spike protein ectodomain and receptor-binding domain IgG correlate with virus neutralization. J Clin Invest, 2020. 130(12): p. 6728–6738.

11. Irsara, C., et al., Clinical validation of the Siemens quantitative SARS-CoV-2 spike IgG assay (sCOVG) reveals improved sensitivity and a good correlation with virus neutralization titers. Clin Chem Lab Med, 2021. 59(8): p. 1453–1462.

12. Padoan, A., et al., Analytical and clinical performances of five immunoassays for the detection of SARS-CoV-2 antibodies in comparison with neutralization activity. EBioMedicine, 2020. 62: p. 103101.

13. Jung, K., et al., Performance evaluation of three automated quantitative immunoassays and their correlation with a surrogate virus neutralization test in coronavirus disease 19 patients and pre-pandemic controls. J Clin Lab Anal, 2021. 35(9): p. e23921.

14. Rubio-Acero, R., et al., In Search of the SARS-CoV-2 Protection Correlate: Head-to-Head Comparison of Two Quantitative S1 Assays in Pre-characterized Oligo-/Asymptomatic Patients. Infect Dis Ther, 2021: p. 1–14.

15. Müller, L., et al., Age-dependent Immune Response to the Biontech/Pfizer BNT162b2 Coronavirus Disease 2019 Vaccination. Clin Infect Dis, 2021. 73(11): p. 2065–2072.

16. Lustig, Y., et al., BNT162b2 COVID-19 vaccine and correlates of humoral immune responses and dynamics: a prospective, single-centre, longitudinal cohort study in health-care workers. Lancet Respir Med, 2021. 9(9): p. 999–1009.

17. Fagni, F., et al., COVID-19 and immune-mediated inflammatory diseases: effect of disease and treatment on COVID-19 outcomes and vaccine responses. Lancet Rheumatol, 2021. 3(10): p. e724–e736.

18. Collier, A.Y., et al., Differential Kinetics of Immune Responses Elicited by Covid-19 Vaccines. N Engl J Med, 2021. 385(21): p. 2010–2012.

19. Munro, A.P.S., et al., Safety and immunogenicity of seven COVID-19 vaccines as a third dose (booster) following two doses of ChAdOx1 nCov-19 or BNT162b2 in the UK (COV-BOOST): a blinded, multicentre, randomised, controlled, phase 2 trial. Lancet, 2021. 398(10318): p. 2258–2276.

20. Collier, D.A., et al., Age-related immune response heterogeneity to SARS-CoV-2 vaccine BNT162b2. Nature, 2021. 596(7872): p. 417–422.

21. Burki, T.K., Omicron variant and booster COVID-19 vaccines. Lancet Respir Med, 2021.

22. Gruell, H., et al., mRNA booster immunization elicits potent neutralizing serum activity against the SARS-CoV-2 Omicron variant. Nat Med, 2022: p. 1–4.

23. Riester, E., et al., Performance evaluation of the Roche Elecsys Anti-SARS-CoV-2 S immunoassay. J Virol Methods, 2021. 297: p. 114271.

24. Muench, P., et al., Development and Validation of the Elecsys Anti-SARS-CoV-2 Immunoassay as a Highly Specific Tool for Determining Past Exposure to SARS-CoV-2. J Clin Microbiol, 2020. 58(10).

25. Jochum, S., et al., Clinical utility of Elecsys Anti-SARS-CoV-2 S assay in COVID-19 vaccination: An exploratory analysis of the mRNA-1273 phase 1 trial. medRxiv, 2021.

26. Chu, L., et al., A preliminary report of a randomized controlled phase 2 trial of the safety and immunogenicity of mRNA-1273 SARS-CoV-2 vaccine. Vaccine, 2021. 39(20): p. 2791–2799.

27. Simel, D.L., G.P. Samsa, and D.B. Matchar, Likelihood ratios with confidence: sample size estimation for diagnostic test studies. J Clin Epidemiol, 1991. 44(8): p. 763–70.

28. R Core Team. R: A language and environment for statistical computing. . R Foundation for Statistical Computing 2020; Available from: https://www.R-project.org/.

29. GmbH, R.D., Elecsys® Anti-SARS-Cov-2 s, Instructions for Use. 2022.

30. Wei, J., et al., Antibody responses to SARS-CoV-2 vaccines in 45,965 adults from the general population of the United Kingdom. Nat Microbiol, 2021. 6(9): p. 1140–1149.

31. Crooke, S.N., et al., Immunosenescence and human vaccine immune responses. Immun Ageing, 2019. 16: p. 25.

32. Schenkelberg, T., Vaccine-induced protection in aging adults and pandemic response. Biochem Biophys Res Commun, 2021. 538: p. 218–220.

33. Zhong, D., et al., Durability of Antibody Levels After Vaccination With mRNA SARS-CoV-2 Vaccine in Individuals With or Without Prior Infection. Jama, 2021.

34. Aziz, N.A., et al., Seroprevalence and correlates of SARS-CoV-2 neutralizing antibodies from a population-based study in Bonn, Germany. Nat Commun, 2021. 12(1): p. 2117.

