## Supplementary material for "Quantifying the vaccine-induced humoral immune response to spike-receptor binding domain as a surrogate for neutralization testing following mRNA-1273 (Spikevax) vaccination against COVID-19"

**Supplementary Table 1. ACOV2S summary statistics, GMC, GMFR, and seroconversion rates in subgroups defined by vaccine dose and age category.** Responder rates are defined as achieving an ACOV2S value ≥0.8 U/mL.

|  | **Day 1** | **Day 15** | **Day 29** | **Day 43** | **Day 57** |
| --- | --- | --- | --- | --- | --- |
| **Placebo** | **n=192** | **n=188** | **n=186** | **n=186** | **n=186** |
| **Median** | 0.400 | 0.400 | 0.400 | 0.400 | 0.400 |
| **Q1–Q3** | 0.400–0.400 | 0.400–0.400 | 0.400–0.400 | 0.400–0.400 | 0.400–0.400 |
| **Min–Max** | 0.400–0.400 | 0.400–0.400 | 0.400–0.400 | 0.400–0.400 | 0.400–0.400 |
| **50 μg** | **n=193** | **n=189** | **n=188** | **n=185** | **n=188** |
| **Median** | 0.400 | 23.4 | 109 | 7356 | 5136 |
| **Q1–Q3** | 0.400–0.400 | 7.06–54.7 | 43.5–196 | 4647–10924 | 3174–7492 |
| **Min–Max** | 0.400–0.400 | 0.400–648 | 3.38–1142 | 1212–25000 | 707–20187 |
| **GMC**  **(95% CI)** | 0.400  (0.400–0.400) | 17.3  (13.5–22.1) | 88.7  (75.9–104) | 6980  (6307–7723) | 4771  (4320–5270) |
| **GSD** | 1.00 | 5.63 | 2.94 | 2.01 | 2.00 |
| **GMFR  (CI)** | 1.00  (1.00–1.00) | 43.1  (33.7–55.3) | 222  (190–259) | 17449  (15769–19308) | 11929  (10800–13175) |
| **Seroconversion rate, % (95% CI)** | 0  (0–1.89) | 92.1  (87.2–95.5) | 100  (98.1–100) | 100  (98.1–100) | 100  (98.1–100) |
| **50 μg,  18‒54 years** | **n=95** | **n=93** | **n=91** | **n=91** | **n=91** |
| **Median** | 0.400 | 35.3 | 136 | 7859 | 5959 |
| **Q1–Q3** | 0.400–0.400 | 11.1–74.3 | 78.1–210 | 5287–12262 | 3684–8120 |
| **Min–Max** | 0.400–0.400 | 0.400–648 | 3.38–1142 | 2062–25000 | 1146–20187 |
| **GMC**  **(95% CI)** | 0.400  (0.400–0.400) | 27.4  (20.0–37.6) | 124  (104–149) | 8143  (7243–9155) | 5577  (4952–6281) |
| **GSD** | 1.00 | 4.63 | 2.38 | 1.76 | 1.77 |
| **GMFR  (CI)** | 1.00  (1.00–1.00) | 68.6  (50.0–94.0) | 311  (260–373) | 20358  (18108–22889) | 13942  (12380–15702) |
| **Seroconversion rate, % (95% CI)** | 0  (0–3.81) | 96.8  (90.9–99.3) | 100  (96.2–100) | 100  (96.2–100) | 100  (96.2–100) |
| **50 μg,  ≥ 55 years** | **n=98** | **n=96** | **n=97** | **n=94** | **n=97** |
| **Median** | 0.400 | 15.3 | 74.9 | 6078 | 4398 |
| **Q1–Q3** | 0.400–0.400 | 3.81–41.5 | 26.6–178 | 3309–10624 | 2175–7143 |
| **Min–Max** | 0.400–0.400 | 0.400–239 | 4.11–768 | 1212–25000 | 707–19511 |
| **GMC**  **(95% CI)** | 0.400  (0.400–0.400) | 11.0  (7.65–15.8) | 64.5  (51.1–81.5) | 6012  (5122–7056) | 4122  (3536–4805) |
| **GSD** | 1.00 | 6.03 | 3.19 | 2.19 | 2.14 |
| **GMFR  (CI)** | 1.00  (1.00–1.00) | 27.5  (19.1–39.6) | 161  (128–204) | 15029  (12804–17641) | 10305  (8840–12013) |
| **Seroconversion rate, % (95% CI)** | 0  (0–3.69) | 87.5  (79.2–93.4) | 100  (96.3–100) | 100  (96.3–100) | 100  (96.3–100) |
| **100 μg** | **n=193** | **n=193** | **n=191** | **n=188** | **n=192** |
| **Median** | 0.400 | 31.9 | 154 | 8484 | 6009 |
| **Q1–Q3** | 0.400**–**0.400 | 8.77–76.0 | 64.7–231 | 5360–13041 | 3375–9693 |
| **Min–Max** | 0.400**–**0.400 | 0.400–633 | 3.14–1426 | 780–25000 | 432–25000 |
| **GMC**  **(95% CI)** | 0.400  (0.400–0.400) | 23.7  (18.5–30.2) | 117  (99.3–137) | 7638  (6862–8502) | 5361  (4793–5995) |
| **GSD** | 1.00 | 5.60 | 3.10 | 2.11 | 2.20 |
| **GMFR  (CI)** | 1.00  (1.00–1.00) | 59.4  (46.5–76.0) | 292  (248–344) | 19068  (17122–21236) | 13390  (11965–14984) |
| **Seroconversion rate, % (95% CI)** | 0  (0–1.89) | 92.7  (88.1–96.0) | 100  (98.1–100) | 100  (98.1–100) | 100  (98.1–100) |
| **100 μg, 18‒54 years** | **n=96** | **n=95** | **n=95** | **n=94** | **n=96** |
| **Median** | 0.400 | 56.7 | 206 | 9249 | 7030 |
| **Q1–Q3** | 0.400**–**0.400 | 21.9–151 | 136–285 | 6750–14354 | 4693–11050 |
| **Min–Max** | 0.400**–**0.400 | 0.400–633 | 10.1–1301 | 1122–25000 | 512–25000 |
| **GMC**  **(95% CI)** | 0.400  (0.400**–**0.400) | 47.5  (35.4–63.8) | 183  (154–217) | 9365  (8273–10602) | 6725  (5888–7680) |
| **GSD** | 1.00 | 4.25 | 2.31 | 1.83 | 1.93 |
| **GMFR  (CI)** | 1.00  (1.00–1.00) | 119  (88.5–160) | 456  (385–541) | 23414  (20682–26506) | 16812  (14721–19199) |
| **Seroconversion rate, % (95% CI)** | 0  (0–3.77) | 97.9  (92.6–99.7) | 100  (96.2–100) | 100  (96.2–100) | 100  (96.2–100) |
| **100 μg, ≥ 55 years** | **n=97** | **n=98** | **n=96** | **n=94** | **n=96** |
| **Median** | 0.400 | 13.0 | 101 | 7014 | 4752 |
| **Q1–Q3** | 0.400**–**0.400 | 3.84–39.7 | 35.2–175 | 3894–10835 | 2586–7772 |
| **Min–Max** | 0.400**–**0.400 | 0.400–500 | 3.14–1426 | 780–25000 | 432–23718 |
| **GMC**  **(95% CI)** | 0.400  (0.400**–**0.400) | 12.1  (8.56–17.0) | 75.0  (58.6–95.8) | 6229  (5272–7361) | 4273  (3603–5068) |
| **GSD** | 1.00 | 5.51 | 3.36 | 2.26 | 2.32 |
| **GMFR  (CI)** | 1.00  (1.00–1.00) | 30.1  (21.3–42.6) | 187  (146–240) | 15495  (13093–18338) | 10638  (8955–12638) |
| **Seroconversion rate, % (95% CI)** | 0  (0–3.73) | 87.6  (79.4–93.4) | 100  (96.3–100) | 100  (96.3–100) | 100  (96.3–100) |
| **GMR**  **(95% CI)** | | | | | |
| **100 vs 50 μg** | 1.00 (1.00–1.00) | 1.37  (0.970–1.94) | 1.32  (1.05–1.65) | 1.09  (0.945–1.27) | 1.12  (0.968–1.30) |
| **18‒54 years: 100 vs 50 μg** | 1.00 (1.00–1.00) | 1.73  (1.13–2.66) | 1.47  (1.15–1.88) | 1.15  (0.971–1.36) | 1.21  (1.01–1.44) |
| **≥ 55 years: 100 vs 50 μg** | 1.00 (1.00–1.00) | 1.09  (0.667–1.80) | 1.16  (0.830–1.63) | 1.04  (0.823–1.30) | 1.04  (0.825–1.30) |
| **100 μg: 18‒54 vs ≥ 55 years** | 1.00 (1.00–1.00) | 3.94  (2.51–6.18) | 2.44  (1.81–3.28) | 1.50  (1.22–1.85) | 1.57  (1.27–1.95) |
| **50 μg: 18‒54 vs ≥ 55 years** | 1.00 (1.00–1.00) | 2.49  (1.54–4.03) | 1.93  (1.43–2.59) | 1.35  (1.11–1.65) | 1.35  (1.11–1.64) |

GMFR, geometric mean fold rise; GMC, geometric mean concentration; GMR, geometric mean ratio; GSD, geometric standard deviation.

**Supplementary Table 2. Summary of qualitative agreement measures between Elecsys ACOV2S and live microneutralization assays (MN_50_ endpoint; reference) with modified ACOV2S cutoffs.**

|  | **0.8 U/mL ACOV2S cutoff** | | **15 U/mL ACOV2S cutoff** | |
| --- | --- | --- | --- | --- |
|  | **Overall database**  **N=2157** | **Excl. Positive ACOV2N participants***  **N=2128** | **Overall database**  **N=2157** | **Excl. Positive ACOV2N participants***  **N=2128** |
| **PPA** | 99.8 (99.2–100) | 99.8 (99.2–100) | 99.3 (98.6–99.8) | 99.4 (98.7–99.8) |
| **NPA** | 92.0 (90.3–93.4) | 91.9 (90.2–93.4) | 93.5 (92.0–94.8) | 93.5 (91.9–94.8) |
| **OPA** | 95.3 (94.3–96.2) | 95.3 (94.3–96.1) | 96.0 (95.1–96.8) | 96.0 (95.1–96.8) |
| **PPV** | 90.3 (88.3–92.0) | 90.1 (88.1–91.9) | 92.0 (90.1–93.6) | 91.8 (89.9–93.5) |
| **NPV** | 99.8 (99.4–100) | 99.8 (99.4–100) | 99.5 (98.9–99.8) | 99.6 (99.0–99.9) |
| **Positive likelihood ratio (95% CI)** | 12.4 (10.3–15.0) | 12.3 (10.2–14.9) | 15.3 (12.4–18.9) | 15.2 (12.3–18.8) |
| **Negative likelihood ratio (95% CI)** | 0.00236  (0.000590–0.00941) | 0.00241 (0.000603–0.00962) | 0.00695 (0.00313–0.0154) | 0.00592 (0.00247–0.0142) |

Data shown as % (95% CI) unless otherwise stated. *All participants with at least one positive ACOV2N result were excluded.
NPA, negative percent agreement; NPV, negative predictive value; OPA, overall percent agreement; PPA, positive percent agreement; PPV, positive predictive value.

**Supplementary Figure 1. Line plots of ACOV2S levels over time in participants with at least one positive ACOV2N result (n=8).** Red horizontal line indicates reactivity cut-off (0.8 U/mL). Dotted grey vertical lines indicate time of vaccination, administered at Days 1 and 29. Red and black dots indicate positive and negative ACOV2N results, respectively.

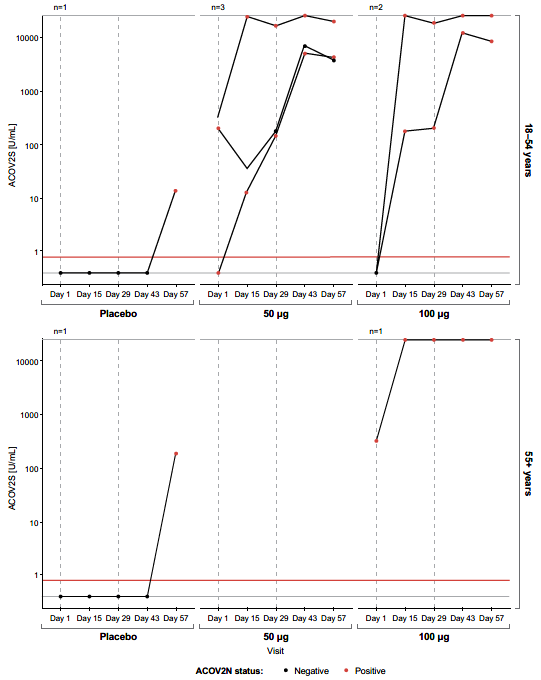

**Supplementary Figure 2. Line plots of ACOV2S levels over time in participants with ACOV2S outlier results (n=6).** Red horizontal line indicates reactivity cut-off (0.8 U/mL). Dotted grey vertical lines indicate time of vaccination, administered at Days 1 and 29.Green and black dots show results that were determined as outliers and non-outliers, respectively.

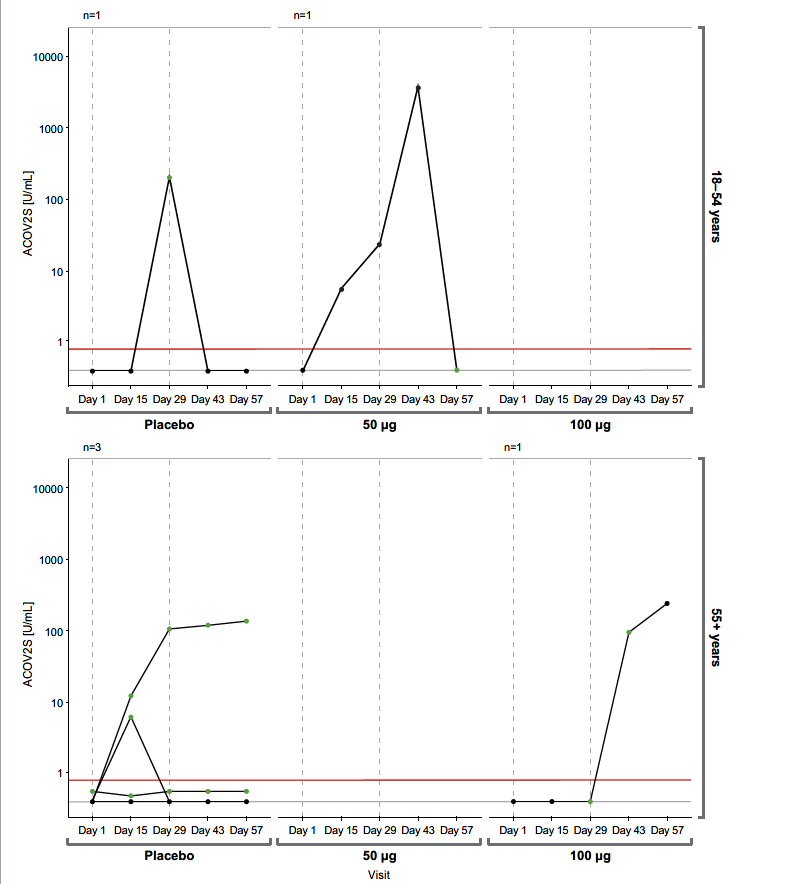

**Supplementary Figure 3. Time course of ACOV2S-measured antibody levels following mRNA-1273 (Spikevax) vaccination in an analysis including ACOV2S outliers.** Antibody levels following vaccination are shown as line plots in Panel A and box plots in Panel B (top and bottom panels showing results from 18–54 and 55+ years age groups, respectively). Dotted grey vertical lines indicate time of vaccination, administered at Days 1 and 29. Box plots show the individual readouts (black dots) and, 25^th^, 50^th^, and 75^th^ percentiles (black box). Red horizontal line indicates reactivity cut-off (0.8 U/mL).

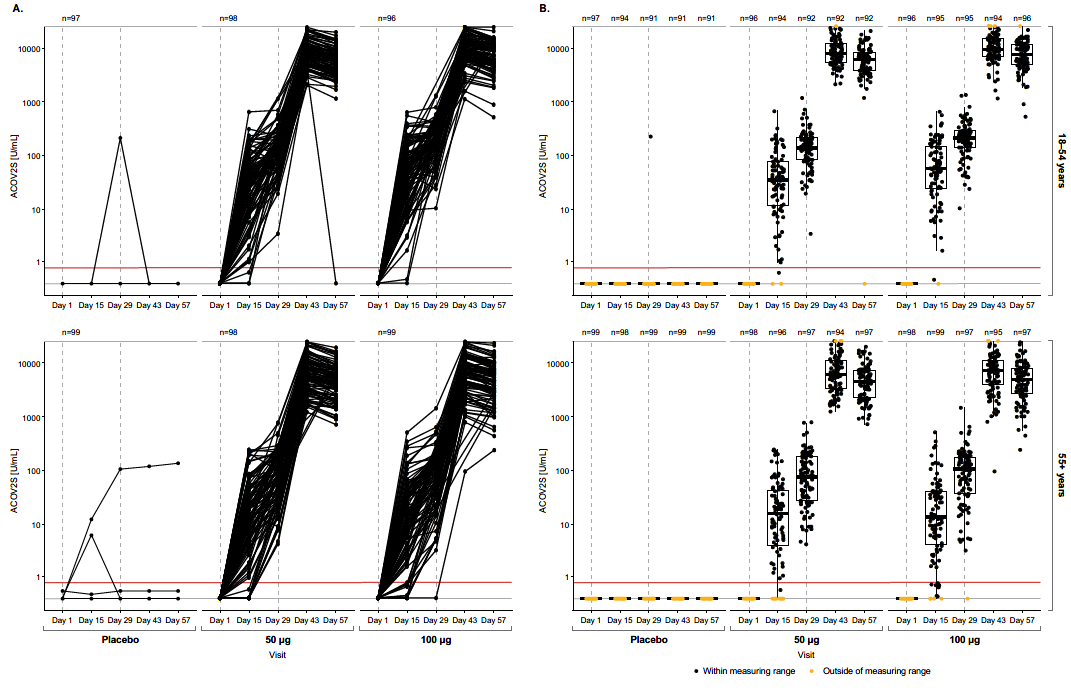

**Supplementary Figure 4 Time-dependent antibody responses as measured by ACOV2S following mRNA-1273 (Spikevax) vaccination in an analysis including ACOV2S outliers.** Reverse cumulative distribution curves allow for comparison of ACOV2S-measured antibody level distributions between subgroups (Panel A) and visits (panel B). Red vertical line indicates reactivity cut off (0.8 U/mL).

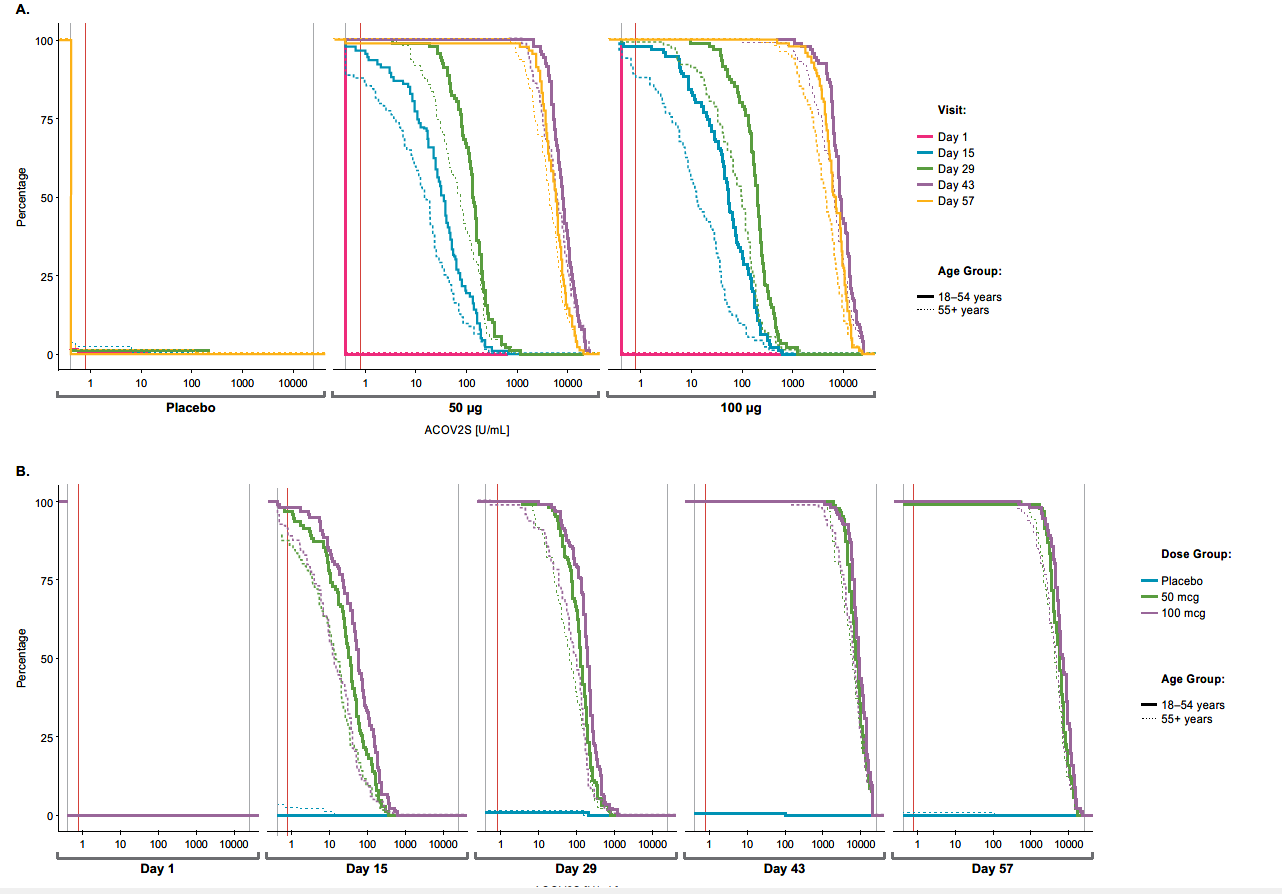

**Supplementary Figure 5. Comparison of ACOV2S and the live microneutralization assay (endpoint MN_50_) for the population including participants with a native SARS-CoV-2 infection.** Scatter plot of ACOV2S vs MN assay results. Data points in red and black are from participants with and without a native SARS-CoV-2 infection, respectively.

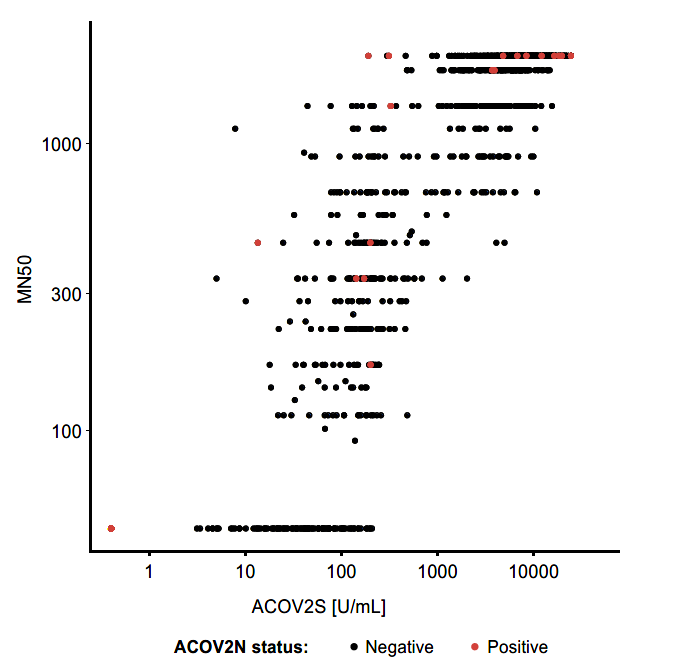
